# Clinical criteria for genetic testing in pediatric oncology show a low specificity and miss every 4^th^ child carrying a cancer predisposition

**DOI:** 10.1101/2022.10.22.22281392

**Authors:** Ulrike Anne Friedrich, Marc Bienias, Claudia Zinke, Maria Prazenicova, Judith Lohse, Arne Jahn, Maria Menzel, Jonas Langanke, Carolin Walter, Rabea Wagener, Triantafyllia Brozou, Julian Varghese, Martin Dugas, Evelin Schröck, Meinolf Suttorp, Arndt Borkhardt, Julia Hauer, Franziska Auer

**Author notes:** shared contribution (Julia Hauer, Franziska Auer).

## Abstract

Clinical checklists are the current gold standard to determine whether a child with cancer shows indications for genetic testing. Nevertheless, the efficacy of these tests to reliably detect genetic cancer predisposition in children with cancer is still insufficiently investigated. Here, we assessed the validity of clinically recognizable signs to identify cancer predisposition by correlating a state-of-the-art clinical checklist to the corresponding whole exome sequencing analysis in an unselected single-center cohort of 139 child-parent datasets. We applied a strict testing to only include autosomal dominant or compound heterozygous cancer-related variants.

Our study reflects a high consent rate for genetic testing (>90%). In total, 1/3^rd^ of patients had a clinical indication for genetic testing according to current recommendations and 10.8% (n=15/139) of children harbored a proven cancer predisposition based on exome sequencing. Out of these only 73.3% (n=11/15) were identified through the clinical checklist. In addition, >2 clinical findings in the applied checklist increased the likelihood to identifying genetic predisposition from 15% to 50%. While our data revealed a high rate of genetic predisposition (50%, n=5/10) in Myelodysplastic Syndrome (MDS) cases, no cancer predisposition variants were identified in the sarcoma and lymphoma group.

In summary, our data showed a low checklist specificity of 68.5%, and missed every 4^th^ child with genetic predisposition. This highlights the drawbacks of sole clinical evaluation to accurately identify all children at risk and underlines the need for routine germline sequencing of pediatric cancers.

## Introduction

Genetic predisposition to pediatric cancer has gained interest and attention over the last decade. What was previously attributed to “chance” or “bad-luck” has now been proven to root in *de novo* occurring, or inherited cancer predispositions in a substantial number of cases. While monogenetic germline variants are currently implicated in the development of up to 10% of all childhood tumors (1-3), the overall impact of genetic predispositions is believed to be much higher, with most yet to be discovered.

The identification of a susceptibility to childhood cancer is highly relevant for the patient, since it may lead to his inclusion in personalized surveillance programs, targeted therapies as well as the option of reproductive counseling and prenatal diagnosis (4). Furthermore, siblings and relatives at risk can subsequently be tested and might benefit from early detection and surveillance opportunities for themselves.

The majority of the currently known classical pediatric cancer predispositions often correlate to a specific clinical phenotype, disease entity, or they occur in a family with a history of cancer (5). This allows the selection of children at high risk who harbor a cancer predisposition, as well as defines which patients should be referred to a human geneticist for further genetic testing (6). Even though some classical cancer predisposition syndromes are easily recognizable (7), others might have an incomplete penetrance or lead to mild phenotypes, which can easily be missed in a daily clinical routine (8). It has already been shown that single clinical indicators, such as the family history are not sufficient to stringently identify patients at risk (9). Therefore, checklists, that combine a multitude of different potential factors, are the current gold standard to judge whether genetic testing in a patient is recommended. These selection tools typically incorporate data concerning family history, type and number of malignancies, as well as specific clinical/physiological features and treatment toxicities (5, 6, 10). Nevertheless, germline genetics is fickle, with many unknowns and blanks that still need to be elucidated. Thus, novel and so far, undiscovered predisposition scenarios, like multigenic susceptibilities that are inherited from healthy parents but synergize in the patient to form a cancer predisposition, might not be recognizable by the phenotypic and clinical data alone (11).

To evaluate the overlap between predisposition selection checklists and detection of cancer predispositions by genetic testing, we correlated both datasets in an unselected cohort of 139 families with pediatric cancer. Here, we show that while the majority of patients harboring a pathogenic or likely pathogenic germline variant are identified by the employed checklist, it occurs at the costs of a high rate of potentially false positives as well as a not negligible percentage of patients that still go unrecognized.

## Results

### Evaluation of 139 children with cancer using a predefined checklist shows a high frequency of indications for genetic testing

The here analyzed cohort consists of 139 families with a child affected from cancer. The cohort includes all cancer entities and was chosen unselected from children treated at the pediatric oncology department at the University Hospital “Carl Gustav Carus” Dresden (Germany) between 2019 and 2021. The recorded data was composed of a detailed clinical and family history of each patient as well as a whole-exome sequencing (WES) analysis of control/healthy tissue of every child as well as (if applicable both of) its parents (**Figure 1A**). The study recruitment showed an overwhelming acceptance rate of around 90% of families consenting to participation. Only a minor fraction of parents (9.2%, n=14/153) refused due to various reasons, including opposition to science, fear, or time restraints (**Figure S1A**). Leukemias and brain tumors comprised the two main subgroups (**Figure 1B, Table 1**). The 33 leukemias could further be subclassified in 22 B-cell acute lymphoblastic leukemias (B-ALL), five T-ALLs and six acute myeloid leukemias (AML) (**Figure S1B**). While the frequency of all included tumor entities reflects the disease distribution within the general pediatric population, the cohort showed an unexpectedly high number of cases with Myelodysplastic Syndrome (MDS) (7.2%, n=10) (**Figure 1B**). Nevertheless, the age at disease onset clustered according to the diagnosis (mean age 7.6 yrs, range 0-18), and the cohort consisted of an equal rate of male and female participants (51.1% vs. 48.9%) (**Figure 1C, Figure S1C-D, Table 1**).

**Table 1:**
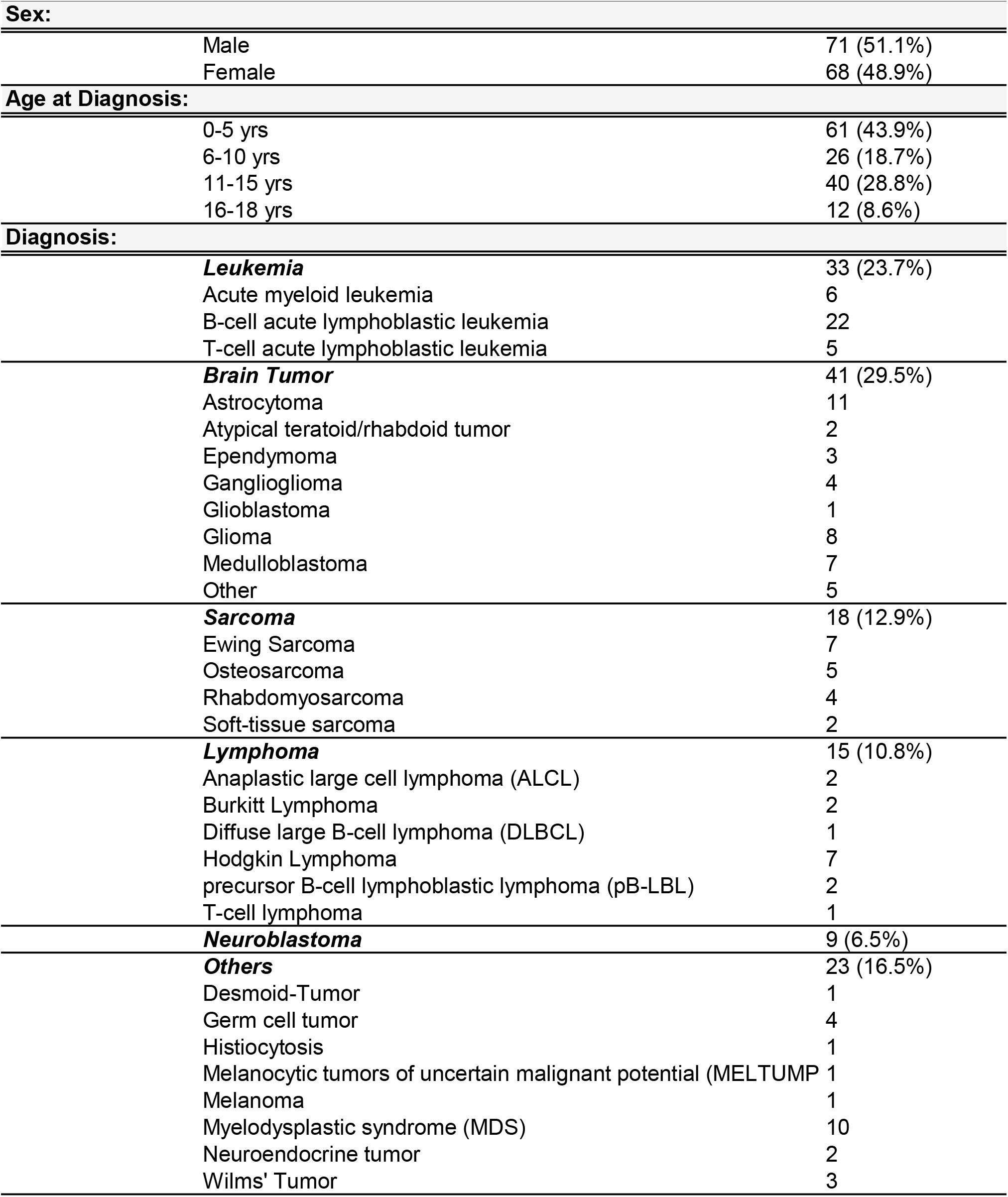
Clinical characteristics of the analyzed cohort (n=139)

| **Sex:** |  |  |
| --- | --- | --- |
|  | Male | 71 (51.1%) |
|  | Female | 68 (48.9%) |
| **Age at Diagnosis:** |  |  |
|  | 0-5 yrs | 61 (43.9%) |
|  | 6-10 yrs | 26 (18.7%) |
|  | 11-15 yrs | 40 (28.8%) |
|  | 16-18 yrs | 12 (8.6%) |
| **Diagnosis:** |  |  |
|  | ***Leukemia*** | 33 (23.7%) |
|  | Acute myeloid leukemia | 6 |
|  | B-cell acute lymphoblastic leukemia | 22 |
|  | T-cell acute lymphoblastic leukemia | 5 |
|  | ***Brain Tumor*** | 41 (29.5%) |
|  | Astrocytoma | 11 |
|  | Atypical teratoid/rhabdoid tumor | 2 |
|  | Ependymoma | 3 |
|  | Ganglioglioma | 4 |
|  | Glioblastoma | 1 |
|  | Glioma | 8 |
|  | Medulloblastoma | 7 |
|  | Other | 5 |
|  | ***Sarcoma*** | 18 (12.9%) |
|  | Ewing Sarcoma | 7 |
|  | Osteosarcoma | 5 |
|  | Rhabdomyosarcoma | 4 |
|  | Soft-tissue sarcoma | 2 |
|  | ***Lymphoma*** | 15 (10.8%) |
|  | Anaplastic large cell lymphoma (ALCL) | 2 |
|  | Burkitt Lymphoma | 2 |
|  | Diffuse large B-cell lymphoma (DLBCL) | 1 |
|  | Hodgkin Lymphoma | 7 |
|  | precursor B-cell lymphoblastic lymphoma (pB-LBL) | 2 |
|  | T-cell lymphoma | 1 |
|  | ***Neuroblastoma*** | 9 (6.5%) |
|  | ***Others*** | 23 (16.5%) |
|  | Desmoid-Tumor | 1 |
|  | Germ cell tumor | 4 |
|  | Histiocytosis | 1 |
|  | Melanocytic tumors of uncertain malignant potential (MELTUMP) | 1 |
|  | Melanoma | 1 |
|  | Myelodysplastic syndrome (MDS) | 10 |
|  | Neuroendocrine tumor | 2 |
|  | Wilms' Tumor | 3 |

**Figure 1:**
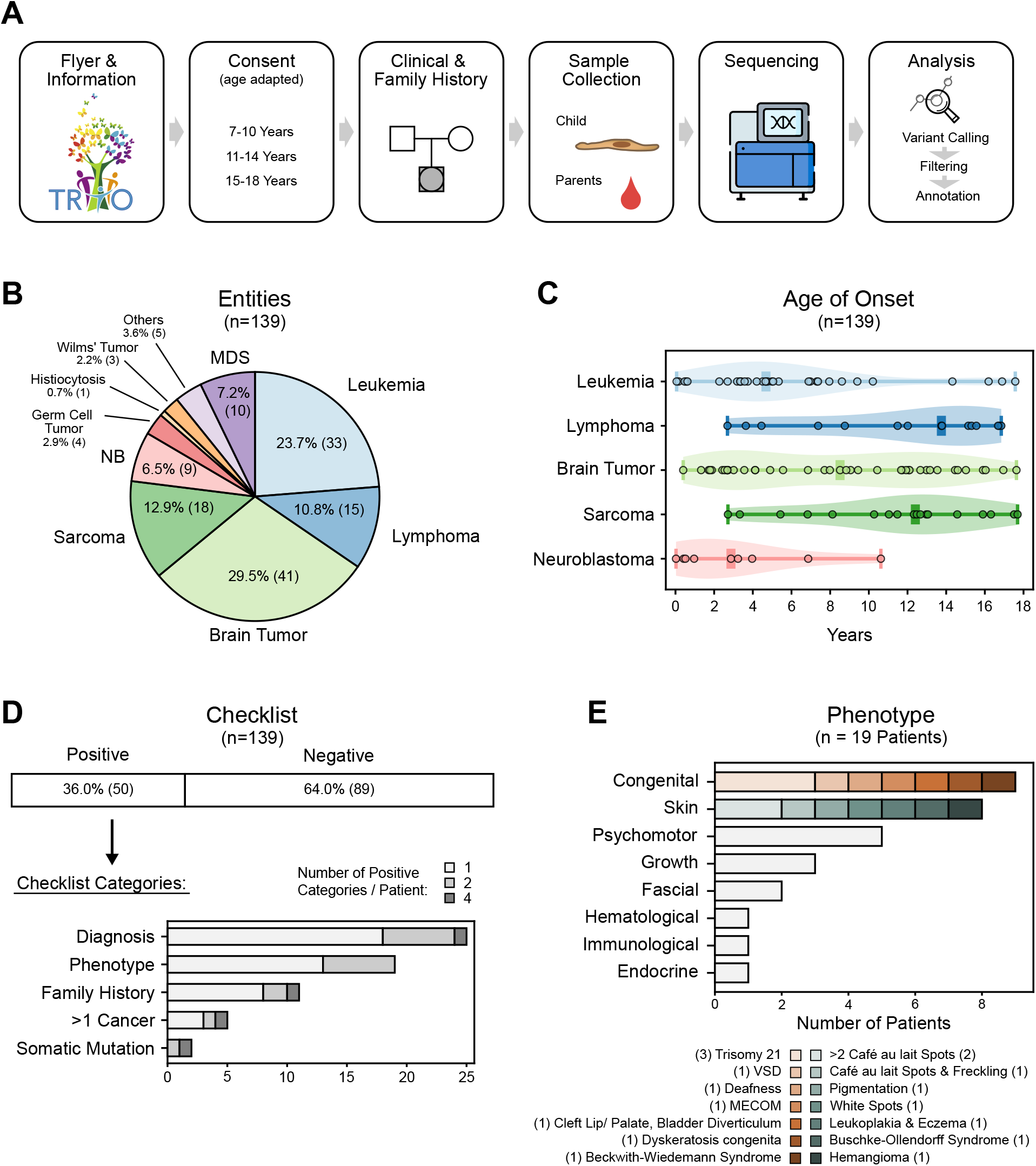
(A) Study workflow comprising the following steps: (1) families are informed and provided with all required information about the study, before (2) signing the study consent. (3) Clinical data, including a three-generation pedigree is collected together with (4) a blood or fibroblast sample. (5) Whole exome sequencing is performed and (6) analyzed to identify all germline variants in the patient. (B) Distribution of tumor entities within the study cohort. Abbreviations: MDS – Myelodysplastic Syndrome, NB – Neuroblastoma. (C) Age distribution of the patients at diagnosis according to disease entity. (D) Patients were examined with respect to the 5 categories of the applied checklist. As soon as one category was positive, the whole questionnaire was considered as positive. Multiple positive categories were possible. (E) Detailed representation of the noted phenotypes within the study cohort. VSD - Ventricular septal defect.

To obtain the overall percentage of pediatric patients with indications for genetic testing, a checklist generated by the German Cancer Society (DKG) and Association for Paediatric Oncology and Haematology (GPOH) (adapted from (5, 6)) was retrospectively applied to evaluate whether a child should be referred for genetic counselling due to the potential of the presence of a pathogenic germline alteration (**Figure S2A**). Subsequently, five main criteria were assessed: type of diagnosis, family history (3-generation pedigree), phenotypic anomalies, total number of malignancies and predisposition indication based on the tumor analysis. The additional criterion of “excessive treatment toxicity” was not considered, as there are no available defined toxicity scoring approaches for pediatric patients across multiple organ systems. According to the checklist, genetic testing for the family should be recommended, as soon as one of the listed identifiers applies. Overall, in our cohort 36% (n=50/139) of children tested positive for an indicative referral to a human geneticist (**Figure 1D**). Most children (n=40/50) were positive for only one criterion and 10 children met two or more criteria. An additional analysis of the most prevalent categories that were marked, showed that the most frequent indicators were the cancer diagnosis (50%) or the additional physical/clinical features (38%) (**Figure 1D**). Particularly for the phenotype, a variety of different anomalies were recorded, with congenital and skin changes being the most frequent (**Figure 1E**). In our cohort, the diagnosis criterion was mainly met through the high amount of MDS patients (40%, n=10/25), while the remaining non-diagnosis-criteria related tumor entities showed a tendency of leukemias and sarcomas towards scoring preferably checklist negative (Fisher’s exact test; 27 vs. 6, p=0.0213; 14 vs. 4, p=0.2924, respectively) (**Figure S2B-C**). A positive family history of cancer was only observed in 22% (**Figure 1D**).

Taken together, the analysis of 139 families with children with cancer through the genetic predisposition checklist evaluated indications for genetic testing in more than 1/3^rd^ of participants (n=50/139), which was mostly based on tumor diagnosis and phenotypic features.

### Exome sequencing shows pathogenic or likely pathogenic variants in 10.8% of the patients

Next, the genetic assessment of cancer predispositions was carried out through a Trio-exome sequencing approach, which comprises WES of the affected child as well as its parents. Altogether, 139 child-parent datasets were analyzed, including 122 Trios (father, mother and child) and 17 Duos (father or mother and child) (**Figure S3**). To objectively screen the pathogenicity of all identified variants, the Cancer Predisposition Sequencing Reporter (CPSR) tool (12), which allows the interpretation of germline variants in the context of cancer predisposition, was used.

Overall, the CPSR analysis yielded a total of 31 pathogenic, 18 likely pathogenic and 1,164 variants of uncertain significance (VUS) within 433 genes across the complete cohort (**Figure 2A, Figure S4A**). While the majority of prioritized pathogenic and likely pathogenic variants were nonsense variants (e.g. splice site, frameshift, stop gained), VUS were mainly comprised of missense variants (**Figure S4B**). However, taking inheritance patterns into account, 21 pathogenic and 11 likely pathogenic variants were excluded from the final results due to known autosomal recessive inheritance (AR) and the lack of compound heterozygosity (**Supplementary Table 1**). Furthermore, 7 variants detected in genes *LEMD3, MLH1, PTPN13, MSH2, COL7A1, EZH2* and *PAX6* were filtered out due a general lack of data regarding the genes’ involvement in cancer predisposition or due to no associations to the specific tumor type presented in the respective child (**Supplementary Table 1**). Lastly, the remaining CPSR classified variants were manually assessed by human geneticists and 10 variants were evaluated as (likely) pathogenic according to ACMG guidelines and further specifications (**Figure 2A**) (13, 14). The data was further complemented with five patients harboring pathogenic variants in autosomal dominant genes *CHEK2, TRNT1, MECOM*, and the autosomal recessive genes *WRAP53* (compound heterozygous) and *ERCC6L2* (biallelic), which were identified apart from the CPSR algorithm during the clinical workup (**Figure 2A**). Thus, the WES analysis within our cohort yielded a total of 15 (likely) pathogenic germline variants in 15 patients (**Figure 2B**). Most of the variants were found in DNA-damage repair pathway related genes, including *BRCA2, ATM, TP53* and *BARD1* or genes involved in proliferative signaling pathways (*NF1* and *TSC1*) (**Figure 2B**). Interestingly, both frameshift variants in *BARD1* were detected in neuroblastoma patients (n=2/9). Also, *POT1* stop variants, one of which was identified in one of the AML patients (15), have recently been linked to myeloid neoplasms (16). Overall, 4 out of 15 variants appeared *de-novo*, while the remaining 11 were inherited from at least one of the parents (**Figure 2B**). Taken together, our analysis yielded a total number of 15 patients with (likely) pathogenic variants connected to cancer predisposition. These were predominantly identified in hematopoietic malignancies (n=8/43) as well as brain tumors (n=5/41) and neuroblastomas (n=2/9), while none were found in lymphomas, sarcomas and the “others” subtype (**Figure 2B**).

**Figure 2:**
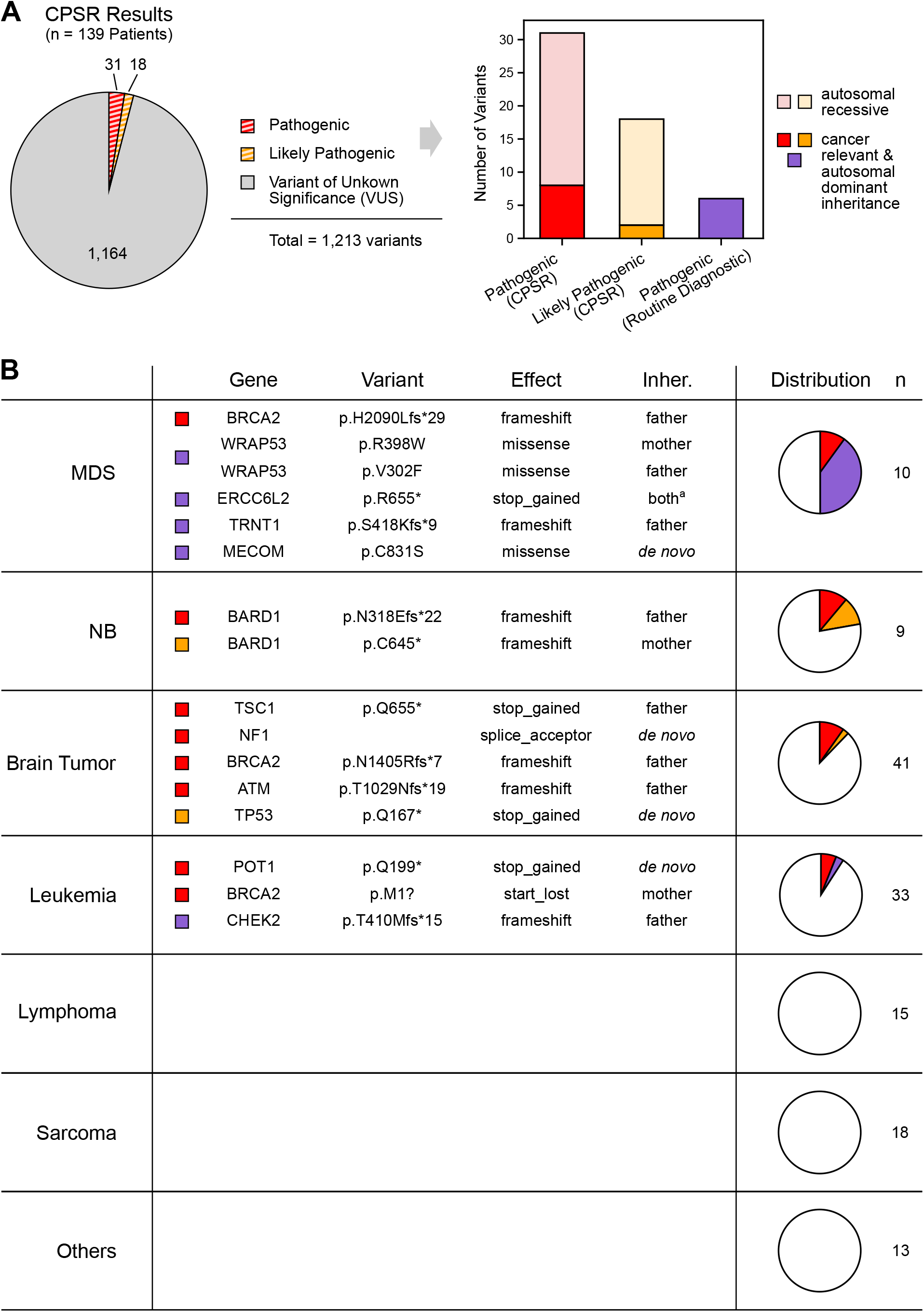
(A) Left: Likely pathogenic and pathogenic variants, as well as variants of uncertain significance (VUS) identified by CPSR within the complete cohort. Right: variants selected for autosomal dominant inheritance and a direct cancer relevance in the respective child are highlighted in darker colouring. Variants displayed in purple represent cancer predispositions, which were detected outside of the CPSR algorithm during clinical workup. (B) List and distribution of selected cancer predisposing variants among different entities in the study cohort; a: the child inherited both variants from the heterozygotic parents and is homozygotic for this variant (biallelic expression); the *CHEK2* variant listed in the leukemia group was found in a patient with transient myeloproliferative disease (TMD). MDS – Myelodysplastic Syndrome, NB – Neuroblastoma, Inher. – Inheritance

### The genetic predisposition checklist shows a high rate of false positives and misses every 4^th^ child carrying a known cancer-causing variant

To address the concordance between the checklist and the detected genomic alterations, both datasets were compared case-by-case. In total, out of 15 patients harboring (likely) pathogenic cancer-related variants, 11 (73.3%) were checklist positive, while four (26.7%) were checklist negative (**Figure 3A, Supplementary Table 2**). There was a higher likelihood to identify a pathogenic variant in patients scoring in multiple criteria, with n=5/10 patients checking multiple criteria and displaying a cancer predisposing variant vs. n=6/40 patients with only one positive criteria, respectively (Fisher’s exact text, p=0.0299; **Figure S5**).

**Figure 3:**
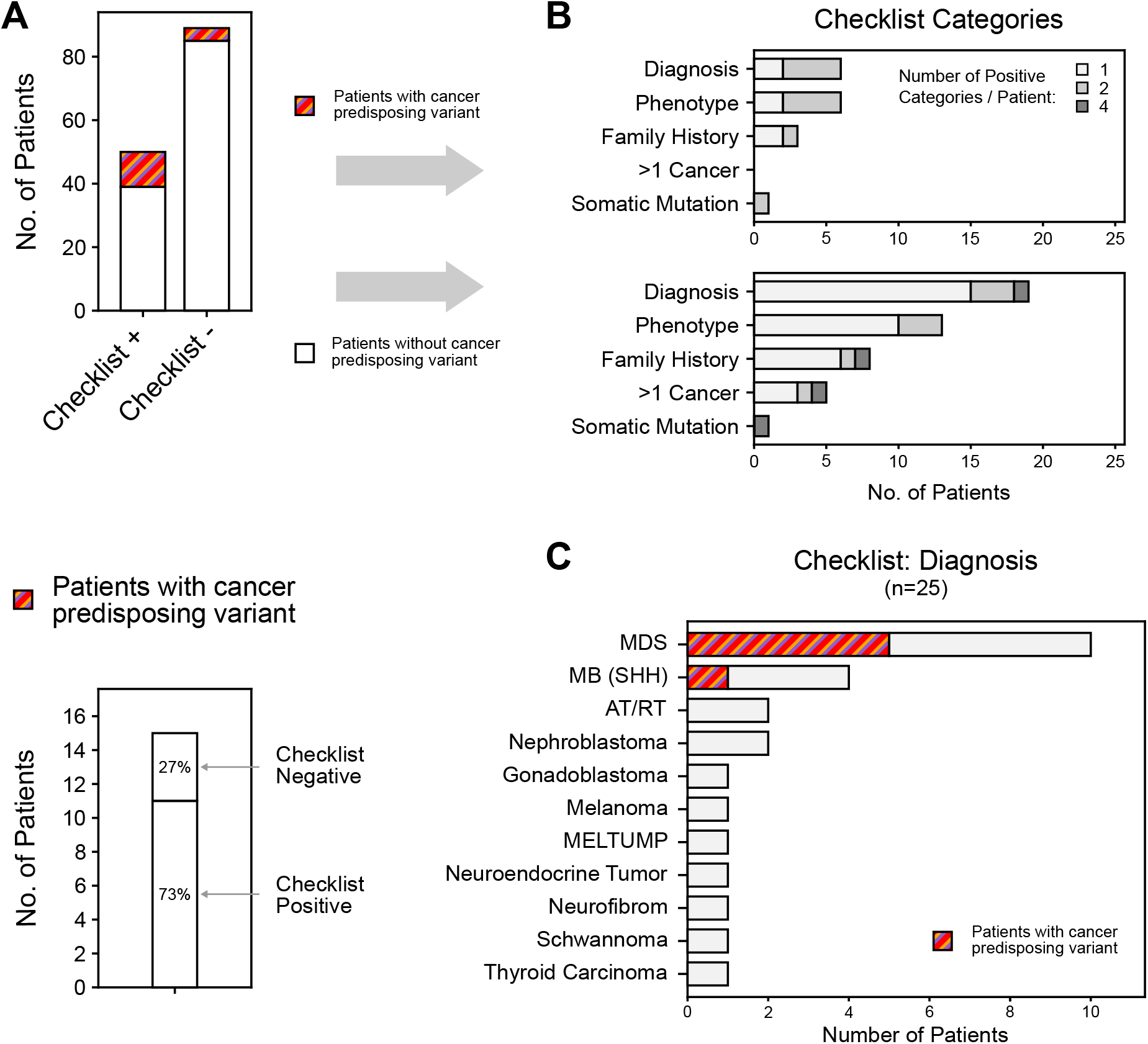
(A) Upper: Distribution of cancer predisposing variants among checklist positive and negative patients. Lower: Patients with predisposing variants grouped for checklist outcome. (B) Checklist positive patients are separated into patients with (top) and without (bottom) cancer predisposing variants. The noted checklist categories are shown for each group individually. Multiple positive categories are possible. (C) Patients which scored checklist positive through the diagnosis criteria. Patients with a proven cancer predisposition are marked accordingly.

Children that were both checklist positive and harbored a (likely) pathogenic variant were almost equally selected via the diagnosis and phenotype criteria, followed by family history (**Figure 3B**). However, both the diagnosis and phenotype criterion scored similarly high in patients with pathogenic germline variants and patients without (**Figure 3B**). Interestingly, 50% of all analyzed MDS patients (n=5/10) carried a pathogenic variant, rendering the diagnosis criterion highly relevant for this specific subtype (**Figure 3C**). By correlating both checklist and genomic data, the clinical evaluation of an underlying cancer predisposition based on state-of-the-art knowledge about genetic predisposition and based on all assessed checklist criteria showed a sensitivity of 73.3% (n=11/15) and specificity of 68.5 % (n=85/124). Taking together, we utilized a very strict analytical approach, focusing on autosomal dominant and compound heterozygous/biallelic cancer-related pathogenic germline variant, which detected a genetic predisposition in every 10-11^th^ child. Out of these, every 4^th^ child would have been missed by clinical selection criteria alone.

## Discussion

Diagnosing germline predisposition in pediatric cancer is essential, as it can convey implications for health care management, treatment and prevention (5). Another aspect is the scientifically often overlooked parent perspective, who might thrive to understand the question as to “why did our child develop cancer?”. Accordingly, our study reports exceedingly high acceptance rates, where 9 out of 10 families willingly participate in germline sequencing. This is also in line with recent meta-studies showing that parents report largely positive attitudes toward childhood genetic testing, appreciating it as beneficial rather than threatening (17). An extended study further confirms that children and young adults (ages 6-21 years) have a basic understanding of inherited disease predisposition within families and that they express an interest to undertake personal genetic testing (18). These analyses also underline the importance of structured, transparent and well-communicated approaches towards genetic testing, paired with a developmentally-sensitive genetic counselling, education and support for the complete family to avoid misinformation and misinterpretation.

As indications for genetic testing in daily practice are easily missed in children (19), user friendly selection tools are useful for pediatric oncologist to identify patients at risk for genetic susceptibilities (6). Nevertheless, the application of the respective checklists in pediatric cancer cohorts shows indications for referral to a clinical geneticist in around every 3^rd^ case, while genetically verified cancer predispositions are only found in up to 15% (9, 20). Here, our data confirm a positive checklist in 36% of all patients within our cohort (n=50/139) and an actual rate of tumor predisposition of 10.8% (n=15/139), out of which 2.9% (n=4/139) scored checklist negative.

While the general frequency per cancer entity in our cohort corresponds to a normal disease distribution, our cohort shows an unusually high rate of MDS patients (7.2%) compared to the expected frequency monitored through the German Childhood Cancer Registry (2.5%) (21). Since MDS is automatically marked as positive for genetic testing in our checklist, this leads to a small bias towards checklist positive cases via the diagnosis criterion in our analysis. Nevertheless, diagnosis still ranks as one of the most applicable categories within our cohort, even after exclusion of surplus MDS cases. Interestingly, overall, an underlying genetic cancer predisposition could be verified in half (n=5) of all analyzed MDS patients, out of which two presented with silent phenotypes. Therefore, our results highlight an exceedingly high rate of genetic predisposition in children with MDS (50%).

Apart from its upsides in referring patients with MDS for genetic testing, the applied checklist only reached a specificity of 68.5%. Thus, based on our current knowledge about cancer predisposition, the majority of checklist positive cases (n=39/50) were categorized as false positives. Nevertheless, the presence of so far unknown cancer related susceptibilities, or of pathogenic variants, which were missed by our analysis, in these patients cannot be ruled out. With new approaches to detect cancer predisposition, like digenic predisposition scenarios or the relevance of structural alterations still being in its infancy (11, 23), the true checklist specificity might indeed be much higher. Also, the high occurrence of VUS in our cohort suggests additional potentially predisposing variants, with novel so far unrecognized cancer susceptibilities and emphasizes the rising need for validated functional testing strategies. This would also explain why some cancer subtypes within our cohort (lymphoma, sarcoma or “others”) showed no pathogenic cancer-related variants. For lymphomas the absence of identified pathogenic variants could be rooted in the application of the utilized CPSR gene list, which might miss immune-deficiency-related genetic predisposition. Also, the sarcoma group includes almost 40% of *EWS-FLI1* driven Ewing Sarcomas. Thus, similar to adult Ewing sarcomas, there might be no additional genetic predisposition relevant (22), or it would only be low penetrant and therefore not detected by our algorithm.

Nevertheless, every 4^th^ patient within our cohort harboring a cancer predisposition would have gone unrecognized by just applying the clinical criteria, which only translates to a checklist sensitivity of 73.3%. While other studies showed higher sensitivity rates of 90-100%, this is mostly attributed to how strictly the respective cancer predisposition is defined (20, 24). Hence, as soon as the search field is broadened outside of the top 5-10% of the classical known cancer predispositions, the sensitivity of clinically implemented checklists drops drastically (9). However, not recognizing a cancer predisposition might be fatal, since its knowledge not only allows surveillance to detect early signs of disease progression but is also crucial to promptly offer hematopoietic stem cell transplantation (HSCT) and choose the right donors, as asymptomatic carriers can occur within a family (25).

Taken together, while our data show that clinical checklists can be helpful to detect genetic predisposition in children with cancer, particular in the context of certain disease entities like MDS, we highlight that a high amount of genetically predisposed children are currently missed. This emphasizes the need for routine clinical germline sequencing in pediatric oncology as an essential necessity to detect currently known predispositions.

## Materials and Methods

### Patient enrollment and data/sample collection

Patients <19 years of age were recruited unselectively at the Pediatric Oncology Department, Dresden (years 2019 until 2021). Consent of the families was obtained according to the Ethical Vote EK 181042019 (Dresden) and in line with the Declaration of Helsinki. The informed consent was obtained by a neutral pediatric oncologist trained in genetic counseling, explaining both benefits and drawbacks of the study. All patients and families were aware that they can retract their consent at any time. After agreeing to participate, the clinical data collection was carried out as described in **Figure 1A**. A checklist provided by the Deutsche Krebsgesellschaft (DKG) and Gesellschaft für pädiatrische Onkologie und Hämatologie (GPOH) Version “checkliste_kio-A1_170509” (2019) was used to evaluate indications for genetic testing. If applicable, DNA of the patient was extracted from cultured skin fibroblasts. Therefore, the skin piece removed during implantation of the central venous catheter, which normally goes to waste, was utilized. For the remaining patients and the parents, peripheral blood (PB) was used.

### Cell culture

Primary fibroblasts were cultured in BIO-AMF™-2 Medium (Biological-Industries, Kibbutz Beit Haemek, Israel) up to 5 passages.

### Sample preparation and analysis pipeline

DNA was extracted from the patient’s fibroblasts using the AllPrep DNA/RNA Mini Kit (Qiagen, Venlo, Netherlands) and from PB of the parents and the remaining patient’s using the QIAamp DNA Blood Mini Kit (Qiagen). Next-generation sequencing libraries for WES were generated using the SureSelect Human All Exon V7 kit (Agilent Technologies, Santa Clara, California, USA). The libraries were sequenced on a NovaSeq 6000 platform (Illumina, San Diego, CA 92122, USA) in paired-end mode (2 × 150bp) and with a final on-target coverage of >100x. Processing of the WES data was carried out as previously described (20) with the following settings: Read files in the fastq format were generated with bcl2fastq v2.19.0, and trimmomatic v0.39 was used to remove adapter and low-quality sequences (26). The alignment to the human reference genome GRCh38 was performed using BWA-MEM v0.7.17 (27) and Samtools v1.2 (28). The tool Peddy 0.4.8 (29) performed gender and relatedness analyses to validate the correct sample assignment and the expected relationship of the patient’s data with the corresponding parents’ data. Single nucleotide variants (SNVs) and insertion/deletions (indels) were called using GATK v4.1.4.1 and VarScan2 v2.3.9 (30), applying the trio mode. Additionally, platypus v0.8.1 (31) was used to call indels (default filter setting). Initial variant interpretation was carried out with the Cancer Predisposition Sequencing Reporter (CPSR, v.1.0.1) (12), which classified the variants as pathogenic, likely pathogenic, variant of uncertain significance (VUS), likely benign or benign. Subsequent genetic assessment according to ACMG and further specifications (see comment above)

### Sanger Sequencing

Validation of the identified pathogenic germline variants was carried out using PCR-based Sanger Sequencing utilizing the following primer pairs:

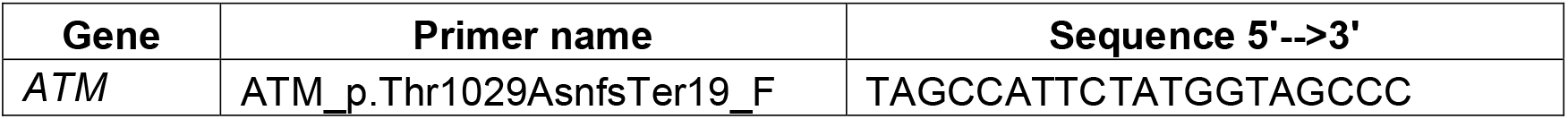
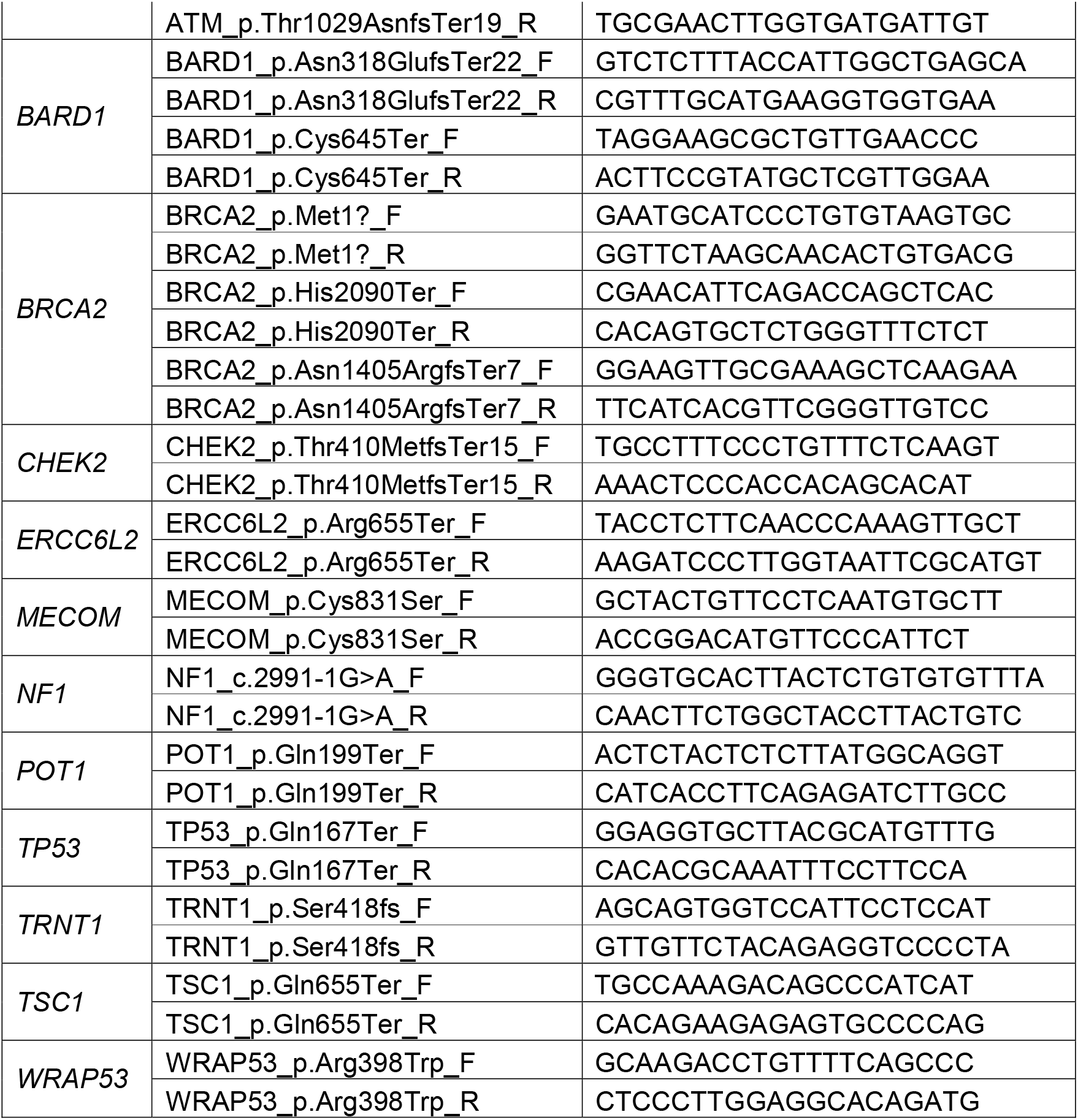

### Data availability statement

(Likely) Pathogenic variant identified in the study were submitted to ClinVar (https://www.ncbi.nlm.nih.gov/clinvar/). The raw datasets generated during and/or analyzed during the current study are not publicly available due to personal data restrictions but are available from the corresponding authors on reasonable request.

## Acknowledgements

The authors would like to thank all members of our groups for useful discussions and for their critical reading of the manuscript. Special thanks go to Carolin Melzig, Anne Schedel, Pia Michler, Rajanya Ghosh and Mina Morcos for sample preparation and documentation.

## References

1. Parsons DW, Roy A, Yang Y, Wang T, Scollon S, Bergstrom K, et al. Diagnostic Yield of Clinical Tumor and Germline Whole-Exome Sequencing for Children With Solid Tumors. JAMA Oncol. 2016;2(5):616–24.

2. Zhang J, Walsh MF, Wu G, Edmonson MN, Gruber TA, Easton J, et al. Germline Mutations in Predisposition Genes in Pediatric Cancer. N Engl J Med. 2015;373(24):2336–46.

3. Narod SA, Stiller C, Lenoir GM. An estimate of the heritable fraction of childhood cancer. Br J Cancer. 1991;63(6):993–9.

4. Brodeur GM, Nichols KE, Plon SE, Schiffman JD, Malkin D. Pediatric Cancer Predisposition and Surveillance: An Overview, and a Tribute to Alfred G. Knudson Jr. Clin Cancer Res. 2017;23(11):e1–e5.

5. Ripperger T, Bielack SS, Borkhardt A, Brecht IB, Burkhardt B, Calaminus G, et al. Childhood cancer predisposition syndromes-A concise review and recommendations by the Cancer Predisposition Working Group of the Society for Pediatric Oncology and Hematology. Am J Med Genet A. 2017;173(4):1017–37.

6. Jongmans MC, Loeffen JL, Waanders E, Hoogerbrugge PM, Ligtenberg MJ, Kuiper RP, et al. Recognition of genetic predisposition in pediatric cancer patients: An easy-to-use selection tool. Eur J Med Genet. 2016;59(3):116–25.

7. Kuhlen M, Borkhardt A. Cancer susceptibility syndromes in children in the area of broad clinical use of massive parallel sequencing. Eur J Pediatr. 2015;174(8):987–97.

8. Knapke S, Nagarajan R, Correll J, Kent D, Burns K. Hereditary cancer risk assessment in a pediatric oncology follow-up clinic. Pediatr Blood Cancer. 2012;58(1):85–9.

9. Byrjalsen A, Hansen TVO, Stoltze UK, Mehrjouy MM, Barnkob NM, Hjalgrim LL, et al. Nationwide germline whole genome sequencing of 198 consecutive pediatric cancer patients reveals a high incidence of cancer prone syndromes. PLoS Genet. 2020;16(12):e1009231.

10. Druker H, Zelley K, McGee RB, Scollon SR, Kohlmann WK, Schneider KA, et al. Genetic Counselor Recommendations for Cancer Predisposition Evaluation and Surveillance in the Pediatric Oncology Patient. Clin Cancer Res. 2017;23(13):e91–e7.

11. Lin M, Nebral K, Gertzen CGW, Ganmore I, Haas OA, Bhatia S, et al. JAK2 p.G571S in B-cell precursor acute lymphoblastic leukemia: a synergizing germline susceptibility. Leukemia. 2019;33(9):2331–5.

12. Nakken S, Saveliev V, Hofmann O, Moller P, Myklebost O, Hovig E. Cancer Predisposition Sequencing Reporter (CPSR): A flexible variant report engine for high-throughput germline screening in cancer. Int J Cancer. 2021;149(11):1955–60.

13. Rehm HL, Berg JS, Brooks LD, Bustamante CD, Evans JP, Landrum MJ, et al. ClinGen--the Clinical Genome Resource. N Engl J Med. 2015;372(23):2235–42.

14. Richards S, Aziz N, Bale S, Bick D, Das S, Gastier-Foster J, et al. Standards and guidelines for the interpretation of sequence variants: a joint consensus recommendation of the American College of Medical Genetics and Genomics and the Association for Molecular Pathology. Genet Med. 2015;17(5):405–24.

15. Michler P, Schedel A, Witschas M, Friedrich UA, Wagener R, Mehtonen J, et al. Germline POT1 Deregulation Can Predispose to Myeloid Malignancies in Childhood. Int J Mol Sci. 2021;22(21).

16. Lim TL, Lieberman DB, Davis AR, Loren AW, Hausler R, Bigdeli A, et al. Germline POT1 variants can predispose to myeloid and lymphoid neoplasms. Leukemia. 2022;36(1):283–7.

17. Lim Q, McGill BC, Quinn VF, Tucker KM, Mizrahi D, Patenaude AF, et al. Parents’ attitudes toward genetic testing of children for health conditions: A systematic review. Clin Genet. 2017;92(6):569–78.

18. McGill BC, Wakefield CE, Vetsch J, Barlow-Stewart K, Kasparian NA, Patenaude AF, et al. Children and young people’s understanding of inherited conditions and their attitudes towards genetic testing: A systematic review. Clin Genet. 2019;95(1):10–22.

19. Merks JH, Caron HN, Hennekam RC. High incidence of malformation syndromes in a series of 1,073 children with cancer. Am J Med Genet A. 2005;134A(2):132–43.

20. Wagener R, Taeubner J, Walter C, Yasin L, Alzoubi D, Bartenhagen C, et al. Comprehensive germline-genomic and clinical profiling in 160 unselected children and adolescents with cancer. Eur J Hum Genet. 2021;29(8):1301–11.

21. Erdmann F, Kaatsch P, Grabow D, Spix C. German Childhood Cancer Registry - Annual Report 2019 (1980-2018). Institute of Medical Biostatistics, Epidemiology and Informatics (IMBEI) at the University Medical Center of the Johannes Gutenberg University Mainz; 2020.

22. Jahn A, Rump A, Widmann TJ, Heining C, Horak P, Hutter B, et al. Comprehensive cancer predisposition testing within the prospective MASTER trial identifies hereditary cancer patients and supports treatment decisions for rare cancers. Ann Oncol. 2022.

23. Sabatella M, Mantere T, Waanders E, Neveling K, Mensenkamp AR, van Dijk F, et al. Optical genome mapping identifies a germline retrotransposon insertion in SMARCB1 in two siblings with atypical teratoid rhabdoid tumors. J Pathol. 2021;255(2):202–11.

24. Chan SH, Chew W, Ishak NDB, Lim WK, Li ST, Tan SH, et al. Clinical relevance of screening checklists for detecting cancer predisposition syndromes in Asian childhood tumours. NPJ Genom Med. 2018;3:30.

25. Kennedy AL, Shimamura A. Genetic predisposition to MDS: clinical features and clonal evolution. Blood. 2019;133(10):1071–85.

26. Bolger AM, Lohse M, Usadel B. Trimmomatic: a flexible trimmer for Illumina sequence data. Bioinformatics. 2014;30(15):2114–20.

27. Li H, Durbin R. Fast and accurate long-read alignment with Burrows-Wheeler transform. Bioinformatics. 2010;26(5):589–95.

28. Li H, Handsaker B, Wysoker A, Fennell T, Ruan J, Homer N, et al. The Sequence Alignment/Map format and SAMtools. Bioinformatics. 2009;25(16):2078–9.

29. Pedersen BS, Quinlan AR. Who’s Who? Detecting and Resolving Sample Anomalies in Human DNA Sequencing Studies with Peddy. Am J Hum Genet. 2017;100(3):406–13.

30. Koboldt DC, Zhang Q, Larson DE, Shen D, McLellan MD, Lin L, et al. VarScan 2: somatic mutation and copy number alteration discovery in cancer by exome sequencing. Genome Res. 2012;22(3):568–76.

31. Rimmer A, Phan H, Mathieson I, Iqbal Z, Twigg SRF, Consortium WGS, et al. Integrating mapping-, assembly- and haplotype-based approaches for calling variants in clinical sequencing applications. Nat Genet. 2014;46(8):912–8.

