## Supplemental Figure and Table Legends for "Clinical criteria for genetic testing in pediatric oncology show a low specificity and miss every 4^th^ child carrying a cancer predisposition"

**Supplementary Figure Legend *Friedrich et al.:***

**Figure S1**:

A) Acceptance of the study among patients of the pediatric oncology department at the University Clinic Dresden. All patients diagnosed between 2019 and 2021 were eligible for study participation.
B) Frequency of the different entity sub-diagnoses among patients in the study cohort, displayed for leukemias, lymphomas and sarcomas.

Abbreviations: AML – acute myeloid leukemia, B-ALL – B-cell acute lymphoblastic leukemia, T-ALL – T-cell acute lymphoblastic leukemia, HL – Hodgkin Lymphoma, ALCL – Anaplastic larger cell lymphoma, Burkitt-L – Burkitt-Lymphoma, DLBCL – Diffuse large B-cell lymphoma, T-LBL – T-lymphoblastic lymphoma, pB-LBL – precursor B-cell lymphoblastic lymphoma, STS – Soft tissue sarcoma, EWS – Ewing sarcoma, OS – Osteosarcoma, RMS – Rhabdomyosarcoma.

C) Age distribution of male and female patients within the study cohort (n=139).
D) Gender distribution within the study cohort.

**Figure S2**:

A) Details of the checklist, which were used to evaluate indications for genetic testing within the study cohort.
B) Numbers of patients per tumor type scoring positive for the Diagnosis criterion. Abbreviations: AT/RT – Atypical teratoid/rhabdoid tumor, MDS – Myelodysplastic Syndrome, MB – Medulloblastoma.
C) Entity distribution of patients separated into questionnaire positive and negative. Abbreviations: MDS – Myelodysplastic Syndrome, NB – Neuroblastoma.

**Figure S3**:

Schematic procedure of variant identification and filtering done for patients participating in the study. Trios include the child and both parents, while duos include the child with only one of its parents. All unfiltered variants are analyzed by the tool Cancer Predisposition Sequencing Reporter (CPSR). The parameters are set to coding only and the following variant categories are further used: Variants of Uncertain Significance (VUS), Likely Pathogenic Variants and Pathogenic Variants. Known technical artefacts have been excluded. The remaining variants were quality controlled according to their relevance to the specific cancer type of the patient and an autosomal dominant or compound heterozygous inheritance. WES – Whole Exome Sequencing.

**Figure S4**:

A) Variants per patient identified by the tool CPSR.
B) The distribution of variant consequence is shown for each variant category.

**Figure S5**:

Patients with and without cancer predisposing variants displayed based on their status of checking one or multiple checklist criteria.

**Supplementary Table 1:**

All Pathogenic and likely pathogenic variants retrieved from the CPSR tool within the complete cohort (n=139). As indicated, autosomal recessive and non-cancer related variants were not further considered in the analysis.

**Supplementary Table 2:**

Genetic and clinical characteristics of patients with pathogenic/likely pathogenic variants (n=15).
