## Supplementary figures and images for "Clinical criteria for genetic testing in pediatric oncology show a low specificity and miss every 4^th^ child carrying a cancer predisposition"

### Supplemental Figures

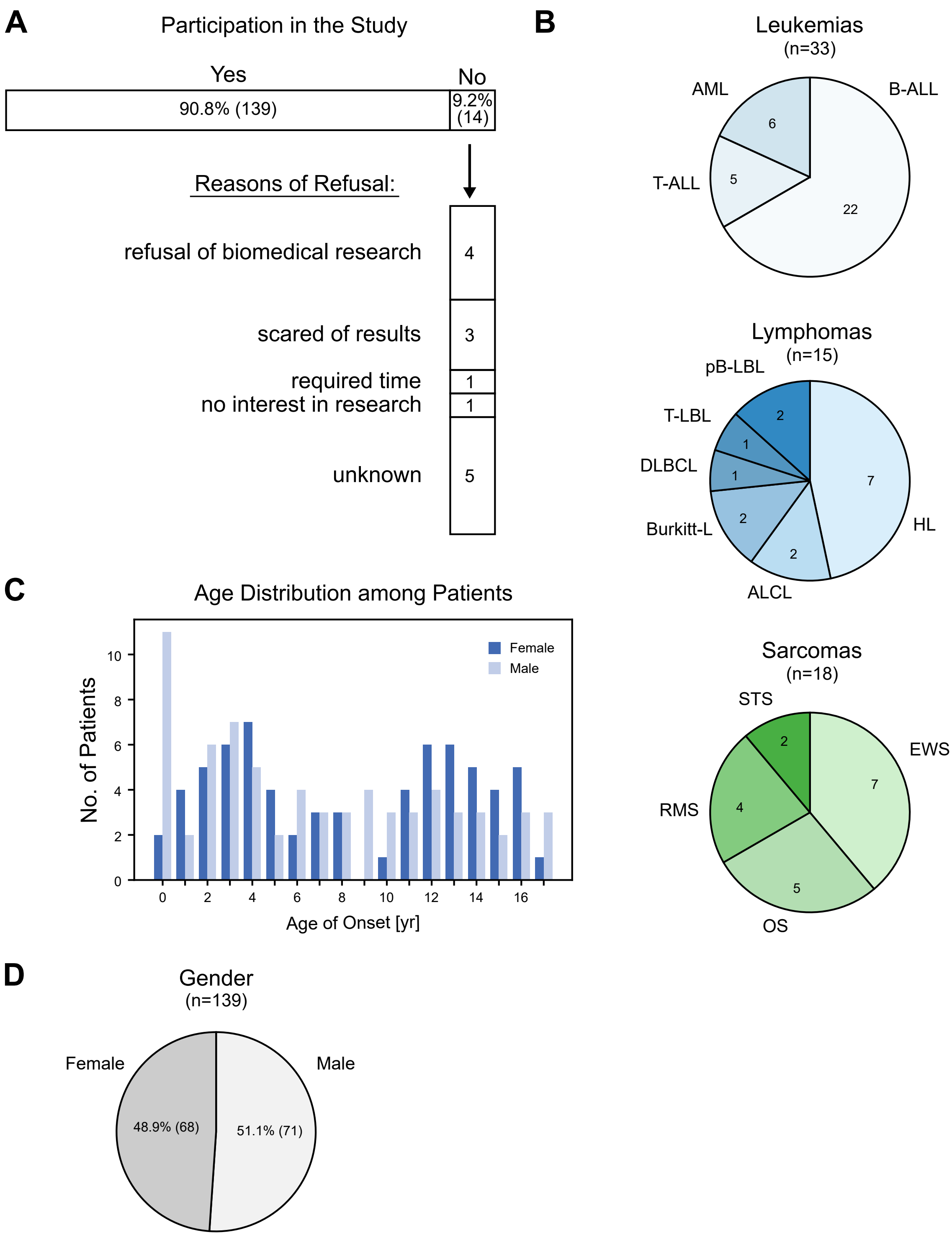

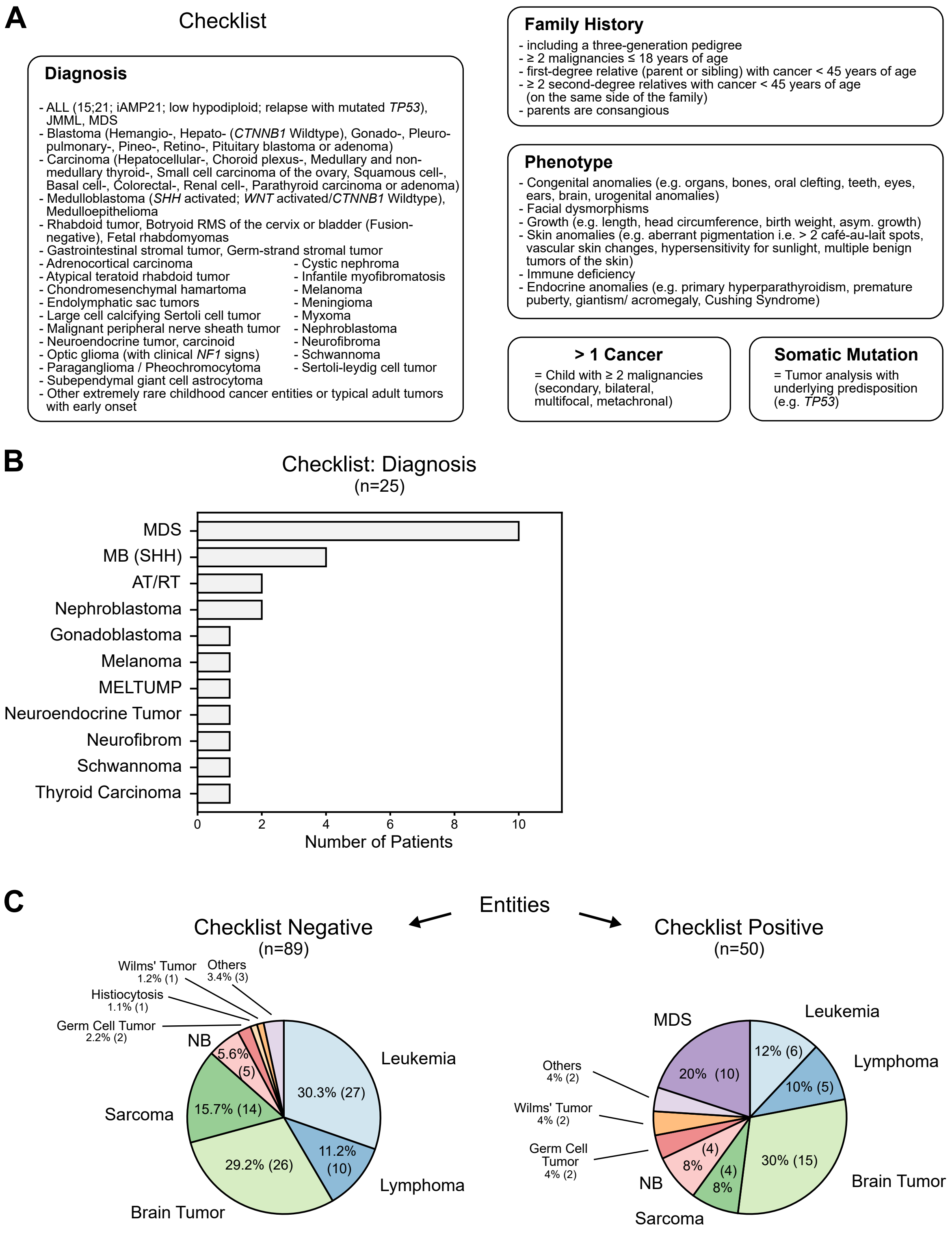

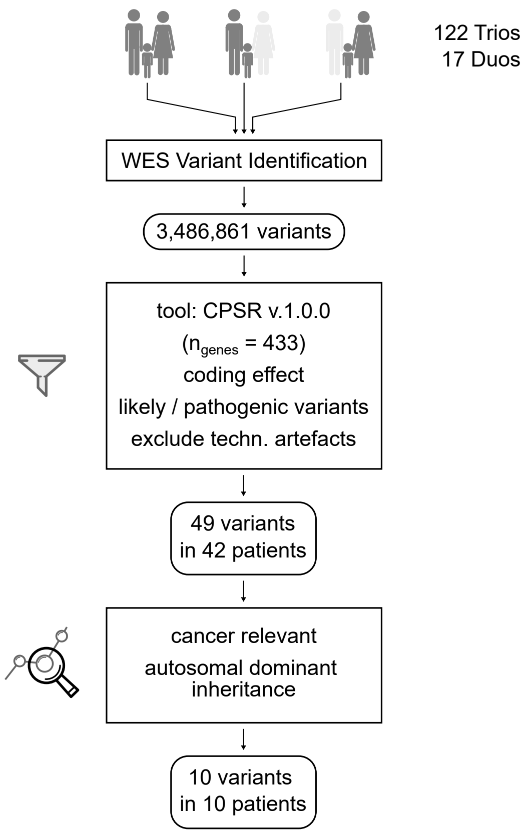

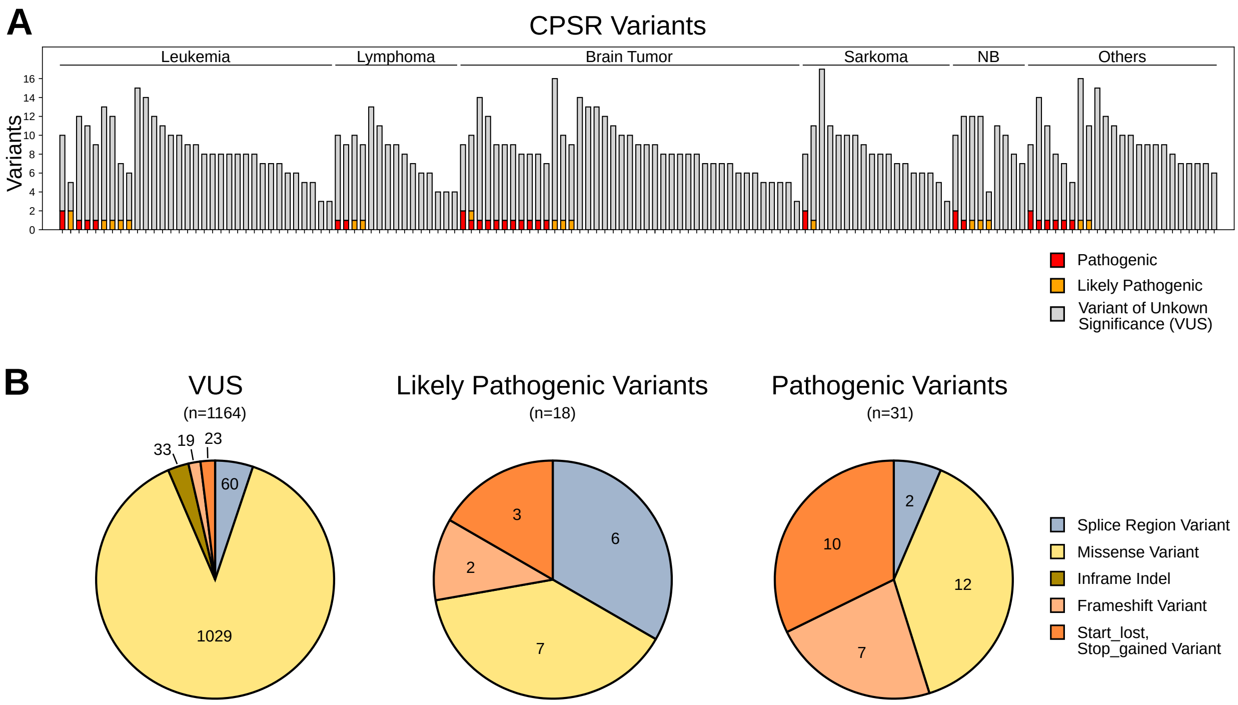

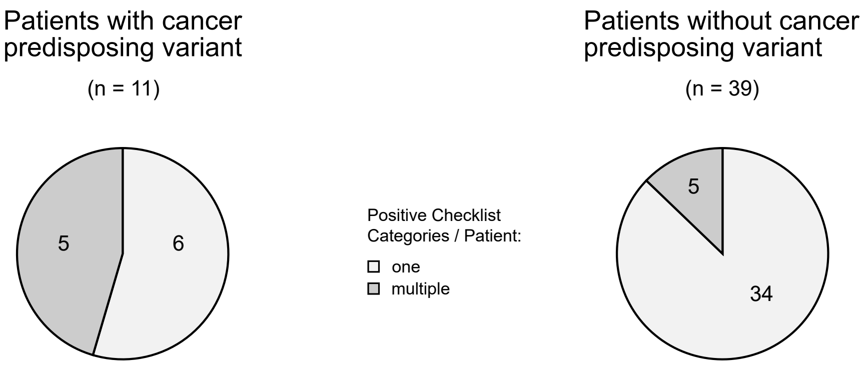
